# On Data-Driven Management of the COVID-19 Outbreak in South Africa

**DOI:** 10.1101/2020.04.07.20057133

**Authors:** Rendani Mbuvha, Tshilidzi Marwala

## Abstract

The rapid spread of the novel coronavirus (SARS-CoV-2) has highlighted the need for the development of rapid mitigating responses under conditions of extreme uncertainty. While numerous works have provided projections of the progression of the pandemic, very little work has been focused on its progression in Africa and South Africa, in particular. In this work, we calibrate the susceptible-infected-recovered (SIR) compartmental model to South African data using initial conditions inferred from progression in Hubei, China and Lombardy, Italy. The results suggest two plausible hypotheses - either the COVID-19 pandemic is still at very early stages of progression in South Africa or a combination of prompt mitigating measures, demographics and social factors have resulted in a slowdown in its spread and severity. We further propose pandemic monitoring and health system capacity metrics for assisting decision-makers in evaluating which of the two hypotheses is most probable.

## 1 Introduction

The novel coronavirus first manifested in the city of Wuhan, China in December 2019. The disease has subsequently spread around the world, leading to the World Health Organisation (WHO) declaring it a pandemic on 11 March 2020 [1]. In South Africa, by 5 April 2020, 1585 people had been confirmed to have infected by the coronavirus with 9 fatalities [2].

African states, together with their counterparts around the globe, have embarked on numerous strict measures to lockdown their countries to “flatten” the curve of COVID-19 cases [3, 4, 5]. The measures are mainly driven by quarantine and isolation strategies that seek to separate the infected population from the susceptible population [3].

These initiatives aim to strategically reduce the surge in infections to a level that their healthcare systems can manage. However, for governments to plan appropriately, they have to answer some of the following questions: how much of the population will be affected? How many will require hospitalisation? When will the country hit the peak infection level? Is the current lockdown effective?

Answers to these questions lie in projections of each country’s infection trajectory. As the pandemic is currently in its early stage in most African countries - calibration of epidemiological models on the basis of available data can prove to be difficult [3]. This effect is escalated by the difference between “confirmed” infected and infected population due to the high number of asymptomatic cases [1] and constrained testing protocols.

In this work, we propose a framework for a data-driven approach to managing the COVID-19 outbreak. We further calibrate the susceptible-infected-recovered (SIR) model based on literature and available data for South Africa. We begin by proposing key metrics for management of a pandemic in section 2, setting out and calibrating the SIR model for South Africa in sections 3 and 3.2. We present our discussion and conclusion in sections 4 and 5.

## 2 Data-Driven Management of the Outbreak

### 2.1 Data and Key Indicators

Effective management of any pandemic will require up to date, good quality data on metrics that are representative of both the spread of the pandemic and healthcare system capacity.

#### 2.1.1 Pandemic Monitoring Metrics

While many jurisdictions report on up to date confirmed case numbers which track the number of individuals who have tested positive for COVID-19, these numbers require to be accompanied by the corresponding number of tests performed for correct interpretation of the progression of the pandemic. This avoids misinterpretation of the trend in tests performed as the trend in prevalence.

An example of this phenomenon is illustrated in figure 1 showing the cumulative confirmed cases, tests performed and positives cases per test in South Africa. It can be seen that the cumulative positive cases reflect the exponential trend in tests performed scaled by the prevalence of the tested cases. The prevalence within the tested population remained relatively stable between 3-4%.

**Figure 1:**
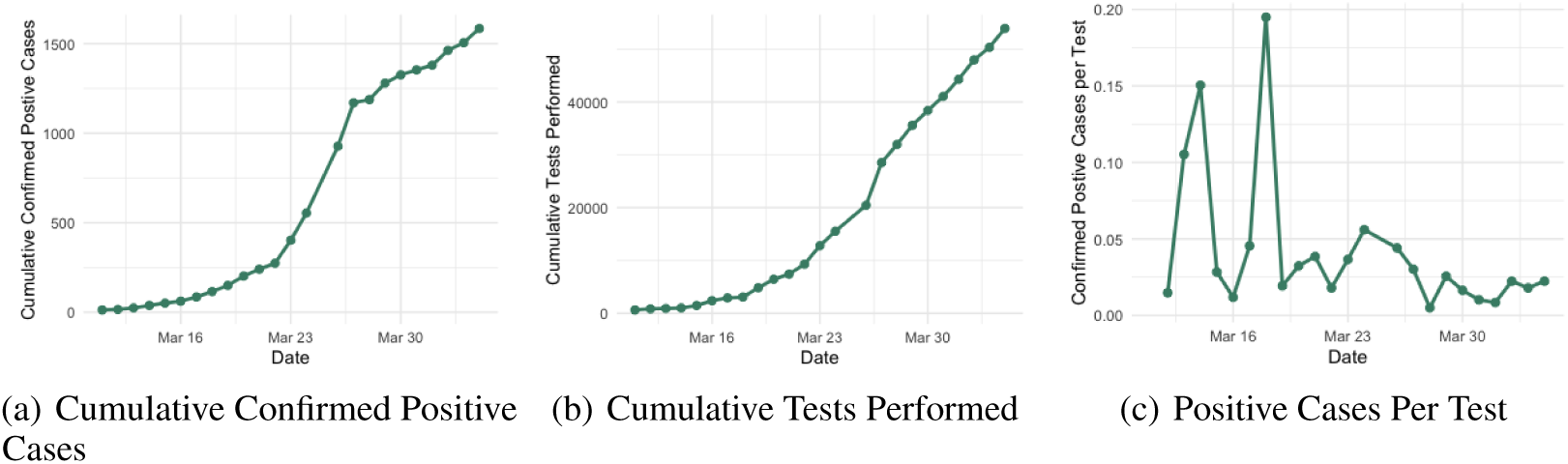
Plots showing the cumulative time series of confirmed positive cases 1(a), tests performed 1(b) and the number of positive cases per test over time. 1(c)

The confirmed positive case metrics are constrained by the healthcare system’s testing capacity and protocols, thus difficult to model. Thus confirmed cases do not necessarily provide an accurate measure of the infected population [3, 5]. Death data, on the other hand, provides a relatively independent unconstrained measure of disease spread. This is because many jurisdictions often have robust protocols for verification of deaths [6].

Other key pandemic spread indicators include:

- The number of persons under investigation (PUIs) at health facilities.
- The numbers of confirmed positive cases per confirmed cases as a result of the contact tracing process.
- The outcomes of mass and randomly sampled screening and testing drives.

#### 2.1.2 Health System Capacity Metrics

The metrics described in 2.1.1 provide the demand side elements of the healthcare resources equation. To co-ordinate an efficient response, States must maintain clean real-time data on their healthcare supply capacity. Such data should include inter-alia some of the following metrics:

- Hospital bed capacity at various levels of care (general, high-care, critical)
- Real-time hospital bed occupation levels
- Stock levels of personal protective equipment (PPE)
- The number of registered healthcare practitioners, both in-service and retired.

States must not be reactive in collecting and curating such data but rather keep databases up to date and ready in pre and post-pandemic times.

## 3 Epidemiological Modelling

Epidemiological Modelling of infectious diseases is dominated by compartmental models which simulate the transition of individuals between various stages of disease [7, 8]. We now introduce the Susceptible-Infected-Recovered (SIR) compartmental model that has been dominant in COVID-19 modelling literature [4, 5].

### 3.1 The Susceptible-Infected-Recovered Model

The SIR is an established epidemiological model for the projection of infectious disease. The SIR models the transition of individuals between three stages of a condition:

- being susceptible to the condition,
- having the condition and being infectious to others and
- having recovered and built immunity for the disease.

The SIR can be interpreted as a three-state Markov chain illustrated diagrammatically in figure 2.

**Figure 2:**
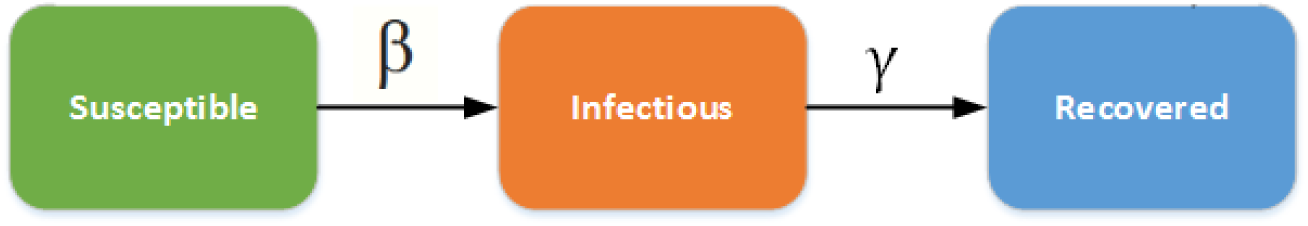
An Illustration of the underlying states of the Susceptible Infected Recovered Model(SIR)

The SIR relies on solving the system of differential equations below representing the analytic trajectory of the infectious disease.

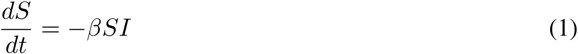

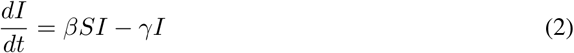

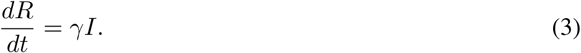

Where S is the susceptible population, I is the infected population, R is the recovered population. *β* is the transmission rate, and *γ* is the recovery rate. 1*/γ*, therefore, becomes the infectious period.

### The Basic Reproductive Number *R*_0_

The contagiousness of a disease is often measured using a metric called the basic reproductive number(*R*_0_). *R*_0_ represents the mean number of additional infections created by one infectious individual in a susceptible population. According to the latest available literature, without accounting for any social distancing the policies the *R*_0_ for COVID-19 between 2 and 2.75[1, 5, 9]. *R*_0_ can be expressed in terms of *γ* and *β* as 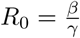.

### 3.2 Localised Calibration

We calibrate the SIR using the reported confirmed South African COVID-19 cases. We follow the framework proposed by [10] for inference of the number of COVID-19 infections from the total number of reported cases to establish initial conditions for the SIR. Specifically, following [10, 11] who suggest that reported confirmed cases represent between 0.2% and 1% of the total infected population. Thus we scale up the confirmed cases reported on 05/04/2020 using correction factors from Hubei, China and Lombardy Italy[10]. We further assume an infectious period of 14 days as suggested by [1] and explore basic reproductive numbers of 2,3,4 inline with related literature [1, 4, 9].

Figure 3 shows the resultant simulated progression of infected cases in South Africa with the above-mentioned parameters.

**Figure 3:**
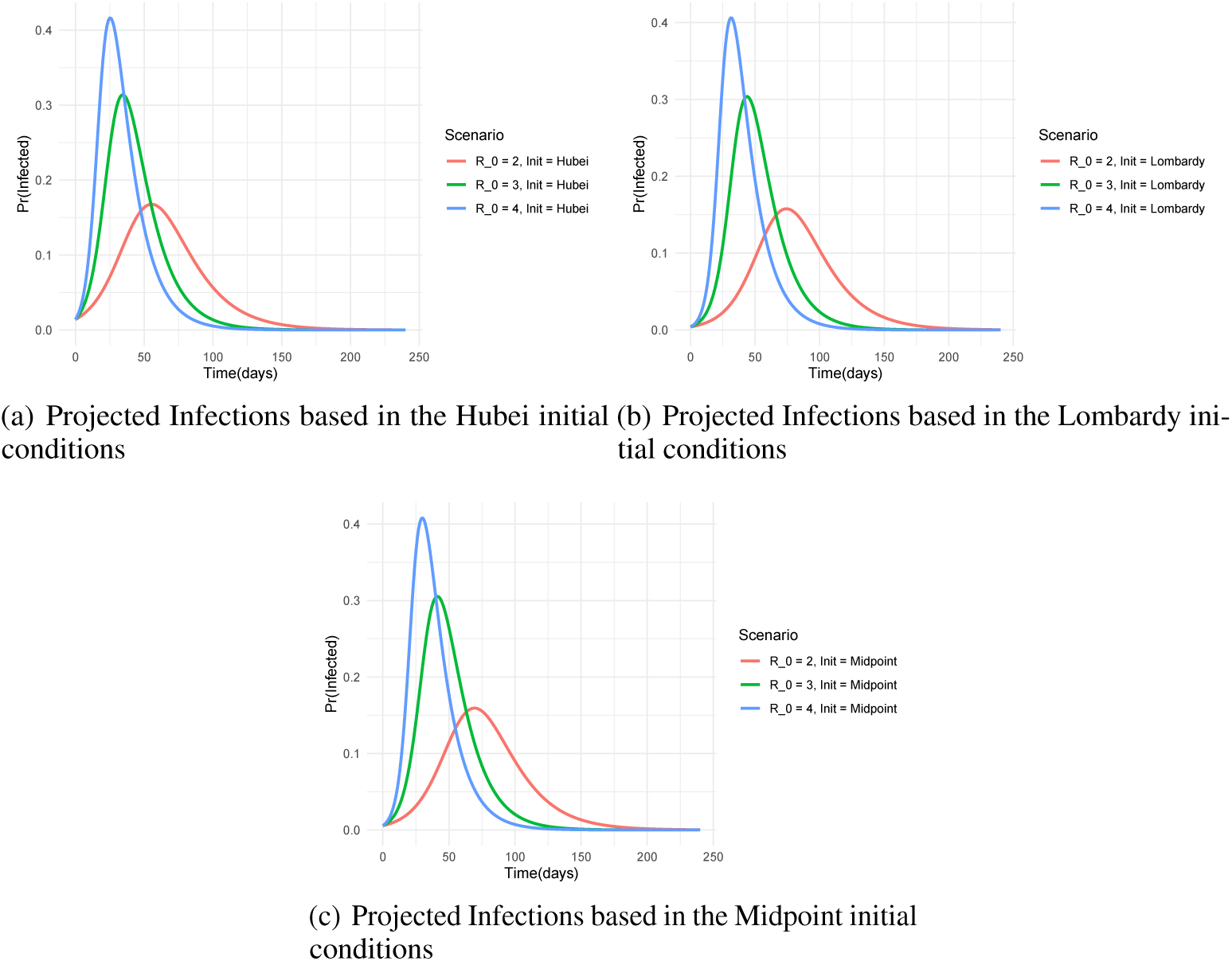
Simulation of COVID-19 infected population progression in South Africa using various initialisation from April 5 2020.

#### 3.2.1 Healthcare Resource Estimation

We use the calibrated SIR model in section 3.2 to estimate the healthcare resources required to manage the pandemic in South Africa. We use the guidance from [1] and [10] as a basis for setting assumptions for transition rates of the infected population into various levels of care.

The healthcare demands of any condition depend on its Clinical Attack Rate (CAR). The CAR is the percentage of the entire population that has been clinically infected (having had mild symptoms, severe or critical) after completion of the outbreak. An empirical study by [10] estimated the of CAR 1% for Wuhan city, about 0.2% for Hubei region and 0.01% throughout China. In Italy, the CAR in Lombardy (the hardest-hit part of Italy) is estimated at 0.7%. Given recent trends in hospital admissions in South Africa, we assume a midpoint CAR of 0.5% for South Africa.

We assume guidance from [1] on the distribution of severity for the clinically attacked individuals such that 80% will display mild or no symptoms, 15% will be severe (General Admission), and 5% will require an Intensive Care Unit (ICU) admission.

### 3.3 Parameter Uncertainty

We have so far assumed that the model parameters of the SIR are fixed albeit at various ranges. In reality there is uncertainty around these parameters that require Bayesian treatment. We illustrate the uncertainty around parameters in figure 5 by sampling *β* and *γ* with midpoint initialisation from log-normal distributions at mean *R*_0_ equivalent to *R*_0_ = 2.5. Future investigations will involve full Bayesian treatment of parameters using Markov Chain Monte Carlo techniques [12, 13]

**Figure 4:**
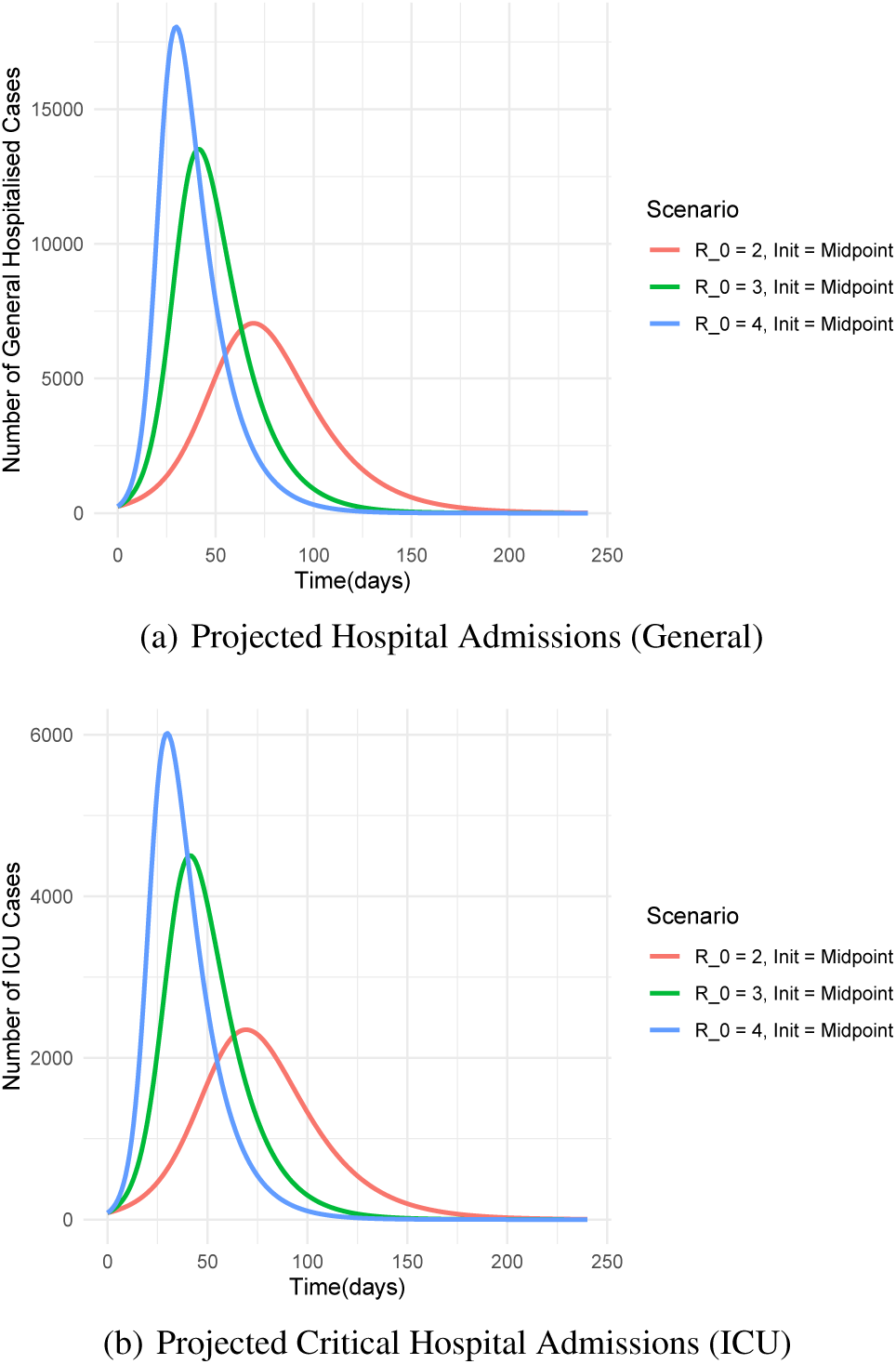
Plots showing the projected hospital admissions based on infection projections in figure 3(c). Figure 4 (a) shows projected general admissions while figure 4 (b) shows projected ICU admissions.

**Figure 5:**
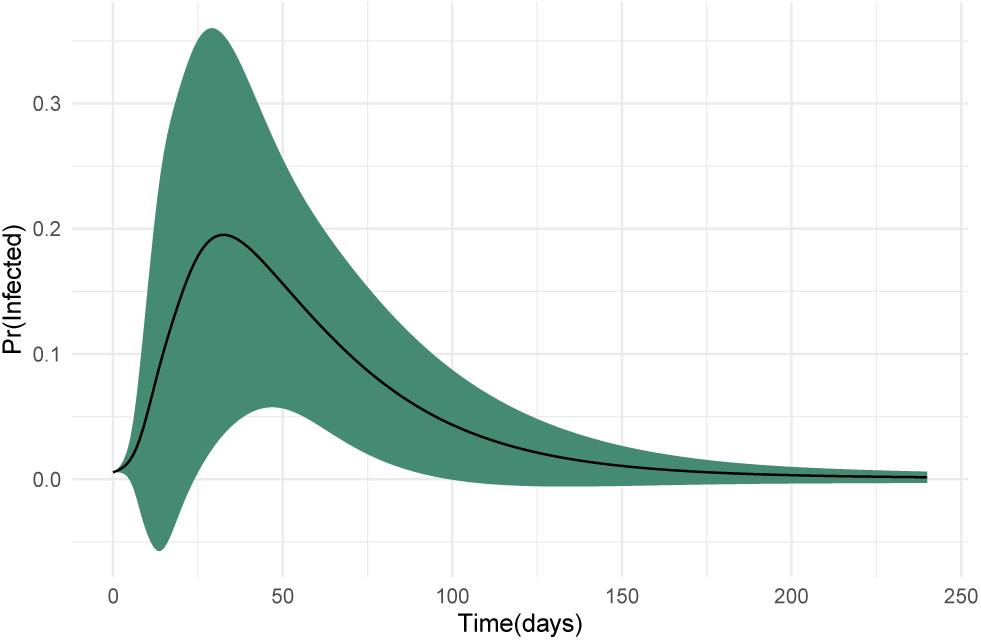
Results of sampling *β* and *γ* from a log-normal distribution with a mean *R*_0_ = 2.5. The green shade shows one standard deviation above and below the mean.

## 4 Discussion

We have attempted to calibrate the SIR model for South Africa and estimate the required healthcare resources using the available literature on inference of infected population. The results show that using the midpoint initial conditions the infected population is expected to peak in: 71 days at *R*_0_ = 2, in 42 days at *R*_0_ = 3 and in 31 days at *R*_0_ = 4. The corresponding estimated general hospital beds required capacities are at 7000 (*R*_0_ = 2), 13500 (*R*_0_ = 3) and 18000 (*R*_0_ = 4). The estimated ICU capacity requirement is estimated at 2350 (*R*_0_ = 2), 4500 (*R*_0_ = 3) and 6000 (*R*_0_ = 4).

The projections provided in this work, however, do not take into account several elements of the South African condition that could significantly alter the dynamics of COVID-19 in South Africa. These fundamental elements include a reduction in imported infections due to the travel ban put in place on 18 March 2020, the subsequent lockdown on 27 March 2020 and a population pyramid skewed towards younger ages relative to Italy and China. Other aggravating conditions unique to South Africa include the high prevalence of the human immunodeficiency viruses (HIV) and densely populated informal urban settlements.

Early indications when contrasting the projections to the current trends in confirmed infections and the limited publicly available hospitalisation data suggest two plausible hypotheses:

1. The pandemic is still in very early stages in South Africa as compared to the rest of the world OR,
2. A combination of factors such as the reduction in imported infections (travel ban), young population (demographic dividend), social dynamics (class separation) and climate have resulted in a significant slowdown in the spread and severity of COVID-19 in South Africa.

A Data-driven pandemic surveillance system including the elements suggested in section 2 is therefore required to evaluate which of the two hypotheses is most likely.

## 5 Conclusion

We have developed a numerical model projection of the COVID-19 infections in South Africa based on publicly available data with initial conditions based on jurisdictions where progression is at advanced stages.

The results suggest that either the progression of COVID-19 in South Africa is still at very early stages or that a combination of prompt mitigating measures, demographics and social factors have resulted in a slowdown in the spread and severity of COVID-19 in South Africa. We further propose data elements that encompass pandemic monitoring metrics and health system capacity metrics to assist decision-makers in evaluating which of the two hypotheses is most likely.

## Data Availability

Publicly Available data

## References

[1] WHO. (2020, feb) Report of the who-china joint mission on coronavirus disease 2019 (covid-19). [Online]. Available: https://www.who.int/docs/default-source/coronaviruse/who-china-joint-mission-on-covid-19-final-report.pdf

[2] V. Marivate, A. de Waal, H. Combrink, O. Lebogo, S. Moodley, N. Mtsweni, V. Rikhotso, J. Welsh, and S. Mkhondwane, “Coronavirus disease (COVID-19) case data - South Africa,” Mar. 2020. [Online]. Available: [https://doi.org/10.5281/zenodo.3732419 (https://doi.org/10.5281/zenodo.3732419)

[3] P. X. Song, L. Wang, Y. Zhou, J. He, B. Zhu, F. Wang, L. Tang, and M. Eisenberg, “An epidemiological forecast model and software assessing interventions on covid-19 epidemic in china,” 2020.

[4] N. Ferguson, D. Laydon, G. Nedjati-Gilani, I. Natsukom, and K. Ainslie. (2020, mar) Report 9: Impact of non-pharmaceutical interventions (npis) to reduce covid-19 mortality and healthcare demand. [Online]. Available: https://www.imperial.ac.uk/media/imperial-college/medicine/mrc-gida/2020-03-16-COVID19-Report-9.pdf

[5] J. Dehning, J. Zierenberg, F. P. Spitzner, M. Wibral, J. P. Neto, M. Wilczek, and V. Priesemann, “Inferring covid-19 spreading rates and potential change points for case number forecasts,” 2020.

[6] T. Jombart, K. van Zandvoort, T. Russell, C. Jarvis, A. Gimma, S. Abbott, S. Clifford, S. Funk, H. Gibbs, Y. Liu, C. Pearson, N. Bosse,, R. M. Eggo, A. J. Kucharski, and J. Edmunds, “Inferring the number of covid-19 cases from recently reported deaths,” 2020.

[7] F. Brauer, Compartmental Models in Epidemiology. Berlin, Heidelberg: Springer Berlin Heidelberg, 2008, pp. 19–79.

[8] J. C. Blackwood and L. M. Childs, “An introduction to compartmental modeling for the budding infectious disease modeler,” Letters in Biomathematics, vol. 5, no. 1, pp. 195–221, 2018. [Online]. Available: https://doi.org/10.1080/23737867.2018.1509026

[9] J. Lourenco, R. Paton, M. Ghafari, M. Kraemer, C. Thompson, P. Simmonds, P. Klenerman, and S. Gupta, “Fundamental principles of epidemic spread highlight the immediate need for large-scale serological surveys to assess the stage of the sars-cov-2 epidemic,” 2020.

[10] Folkhälsomyndigheten, “Skattning av behov av slutenvårdsplatser covid-19 (den 20 mars 2020, uppdaterad 27 mars 2020),” mar 2020. [Online]. Available: https://www.folkhalsomyndigheten.se/nyheter-och-press/nyhetsarkiv/2020/mars/nya-berakningar-av-hur-manga-vardplatser-som-behovs-pa-grund-av-covid-19/

[11] I. Covid, C. J. Murray et al., “Forecasting covid-19 impact on hospital bed-days, icu-days, ventilator-days and deaths by us state in the next 4 months,” medRxiv, 2020.

[12] I. Boulkaibet, T. Marwala, L. Mthembu, M. I. Friswell, and S. Adhikari, “Sampling techniques in bayesian finite element model updating,” in Topics in Model Validation and Uncertainty Quantification, Volume 4, T. Simmermacher, S. Cogan, L. Horta, and R. Barthorpe, Eds. New York, NY: Springer New York, 2012, pp. 75–83.

[13] R. Mbuvha, I. Boulkaibet, and T. Marwala, “Bayesian automatic relevance determination for feature selection in credit default modelling,” in International Conference on Artificial Neural Networks. Springer, 2019, pp. 420–425.

